# EPIDEMIOLOGICAL AND CLINICAL CHARACTERISTICS OF COVID-19 PATIENTS IN KENYA

**DOI:** 10.1101/2020.11.09.20228106

**Authors:** Loice Achieng Ombajo, Nyamai Mutono, Paul Sudi, Mbuvi Mutua, Mohammed Sood, Alliy Muhammad Ali Loo, Phoebe Juma, Jackline Odhiambo, Reena Shah, Frederick Wangai, Marybeth Maritim, Omu Anzala, Patrick Amoth, Evans Kamuri, Waweru Munyu, SM Thumbi

## Abstract

**Background:** More than 49,000 cases of infection and 900 deaths from COVID-19 have been recorded in the Kenya. However, the characteristics and risk factors for severe outcomes among hospitalized COVID-19 patients in this setting have not been described.

**Methods:** We extracted demographic, laboratory, clinical and outcome data from medical records of RT-PCR confirmed SARS-CoV2 patients admitted in six hospitals in Kenya between March and September, 2020. We used Cox proportional hazards regressions to determine factors related to in-hospital mortality.

**Results:** Data from 787 COVID-19 patients was available. The median age was 43 years (IQR 30-53), with 505 (64%) males. At admission, 455 (58%) were symptomatic. The commonest symptoms were cough (337, 43%), loss of taste or smell (279, 35%), and fever (126, 16%). Co-morbidities were reported in 340 (43%), with cardiovascular disease, diabetes and HIV documented in 130 (17%), 116 (15%), 53 (7%) respectively. 90 (11%) were admitted to ICU for a mean of 11 days, 52 (7%) were ventilated with a mean of 10 days, 107 (14%) died. The risk of death increased with age [hazard ratio (HR) 1.57 (95% CI 1.13 – 2.19)] for persons >60 years compared to those <60 years old; having co-morbidities [HR 2.34 (1.68 – 3.25)]; and among males [HR 1.76 (1.27, 2.44)] compared to females. Elevated white blood cell count and aspartate aminotransferase were associated with higher risk of death.

**Conclusions:** We identify the risk factors for mortality that may guide stratification of high risk patients.

## Introduction

Ten months since the first case of coronavirus disease (COVID-19) was reported, more than 45 million cases and 1.2 million people have died from the disease globally. Africa has recorded 1.8 million cases and 42,000 deaths with South Africa reporting the most cases on the continent [1]. Kenya reported its first case on 13th March 2020 and has recorded more than 45,000 cases and 1,000 COVID-19 deaths within 8 months with established community transmission in all the 47 counties as documented by the Ministry of Health Emergency Operations Centre.

The epidemiological and clinical characteristics of COVID 19 patients are not fully known. Initial data from China reported a median age of 47 in patients with COVID-19, majority of patients were male with only 5% requiring ICU care and a 1.4% mortality [2]. On the other hand, limited data from Africa have reported higher mortality for <20 years compared to 20-39 year olds [3], which differs significantly from what is reported elsewhere and underscores the need to understand the disease dynamics in multiple settings.

The COVID-19 disease spectrum ranges from asymptomatic, mild, moderate, severe to critical disease [4]. About 5% of patients have critical disease which is defined as respiratory failure, shock or multiorgan dysfunction, usually exacerbated by immune hyperactivation such as the cytokine storm [5]. The most common laboratory findings include lymphopenia, elevated aminotransferases, elevated lactate dehydrogenase, elevated C reactive protein (CRP) and elevated D-dimer levels [6]. Common chest radiological findings include ground glass opacities in 83%, mixed ground glass opacities and consolidation in 58%, pleural thickening in 50% and interlobular septal thickening in 48% [7].

The overall COVID-19 case fatality rate approximates 2.3% [5]. Most fatalities are patients with advanced age or underlying co-morbidities. Case fatality also varies by region depending on population characteristics, for example Italy which has an older population reported a case fatality rate of 7.2% as compared to Korea where the median age is 40 years and the case fatality rate was 0.7%[8,9]. In an analysis of 300,000 patients with confirmed COVID-19 in the United States, mortality was twelve times higher among patients with co-morbidities [10]. Co-morbidities shown to be risk factors for severe illness include cardiovascular disease, smoking, diabetes mellitus, hypertension, chronic lung disease, chronic kidney disease, and obesity [5]. Some laboratory findings have been also associated with poor outcomes. These include lymphopenia, thrombocytopenia, deranged liver function tests, elevated lactate dehydrogenase and raised inflammatory markers [11].

Whereas detailed reports of clinical features and outcome of patients hospitalised with COVID-19 are increasing from various parts of the world, data from Africa is scarce. In this paper we report the epidemiologic and clinical features of patients admitted with COVID-19 to Kenyan Hospitals and describe the risk factors for mortality.

## Methods

### Study design

This multi-center cohort study recruited patients admitted into six hospitals with laboratory confirmed diagnosis of SARS-CoV2 between 14^th^ March 2020 and 17^th^ September 2020. The six hospitals (3 public hospitals and 3 private hospitals) had provided dedicated facilities with isolation beds for COVID-19 patients. Two of the public hospitals (Kenyatta National Referral Hospital and Mbagathi Hospital) are located in Nairobi while one public hospital (Coast General Teaching and Referral Hospital) is in Mombasa County at the Coast. The three private hospitals (Nairobi Hospital, Aga Khan University Hospital, Avenue Hospital) are located in Nairobi.

Following the report of the first SARS-CoV-2 case in Kenya on 13^th^ March 2020, the government policy required isolation of all infected persons, including those that were asymptomatic, in health-facilities, prior to the adoption of home-based care guidelines on 12^th^ June 2020. COVID-19 patients of all ages were recruited into the study. Confirmation of SARS-CoV-2 infection was through real-time PCR testing of nasal and oral-pharyngeal swabs at the government designated COVID-19 testing centres. The study received ethical approval from the Kenyatta National Hospital-University of Nairobi Ethics and Research Committee (approval number P223/03/2020).

### Procedures

We developed a detailed questionnaire that was used to systematically extract information from the medical records of COVID-19 patients admitted into these study hospitals. Briefly, the questionnaires captured patient information at admission into the hospital, during the hospitalization until discharge or death. Medical records of the SARS-CoV2 positive patients admitted in the study hospitals were reviewed by a team of trained physicians. They extracted data including patient demographic data, medical history, underlying co-morbidities, clinical symptoms, laboratory findings, management and treatment measures and outcome data. Presenting symptoms at admission and during the hospital stay were obtained.

All patients had daily pulse oximetry and blood samples drawn for blood count, liver and renal function tests within 24 hours of admission. Patients who had moderate to severe illness got a chest radiograph, CRP and D-dimers in facilities where these investigations were available. Other investigations were ordered as informed by the clinical scenario. Patient with low oxygen saturation were given supplemental oxygen via nasal prongs or masks as appropriate. Patients requiring further respiratory support were admitted to the ICU for either non-invasive or invasive ventilation.

At the beginning of the outbreak in Kenya, patients were given supportive care, many patients at the time also received various repurposed drugs including azithromycin and hydroxychloroquine, in keeping with the little evidence available at the time. As more data became available, dexamethasone became standard of care for severely ill patients on oxygen or requiring mechanical ventilaion and the use of hydroxychloroquine and azithromycin was abandoned. Access to novel antivirals is still poor in the country. The duration from onset of symptoms to hospital admission, to requiring ventilation, to ICU admission and to death were recorded.

### Data analysis

Categorical variables were presented as counts and percentages, and continuous variables as means with standard deviation for normally distributed data, or median with inter-quantile ranges for data that were not normally distributed. Independent group *t-*tests were used to compare means of continuous variables for normally distributed data, and the non-parametric Mann-Whitney U test for the data not normally distributed. Comparison between proportions of categorical variables was done using the Chi-square tests. The data from laboratory tests was categorized as normal (within the normal range) or abnormal (outside the normal range).

For survival analysis, the primary outcome of this study was COVID-19 related death, defined as death among patients admitted into the hospital with RT-PCR confirmed SARS-CoV-2 infection and complications associated with COVID-19. Cox proportional hazards regressions were used to calculate Hazard ratios (HRs) associated with the demographic, underlying co-morbidities, symptoms, clinical and laboratory characteristics were calculated, and the confidence intervals set at 95%. All the statistical analysis was carried out using the R statistical software [12].

## Results

### Baseline characteristics of the study patients

A total of 787 patients from six health facilities were recruited into the study. The median age was 43 years, with 42% of the patients being below 40 years. Majority of the patients (64%) were male. Nearly two thirds (67%) of the patients had visited the health facility while the rest had been admitted following the initial government regulations of isolating all laboratory confirmed positive cases in health facilities. Of the admitted patients, 43% had underlying conditions, with the most common being cardiovascular diseases (17%), diabetes (15%), HIV (7%), cancer (4%), chronic renal disease (3%) and chronic obstructive pulmonary disease (3%) (Table 1).

**Table 1:** **Demographics and baseline characteristics of the patients admitted in Kenyan health facilities with COVID-19**

| Parameter | All patients<br>(n=787) | Survivors<br>(n=680) | Non-survivors<br>(107) | p-value |
| --- | --- | --- | --- | --- |
| *Age in years* median (SD) | 43 (0 - 109) | 41 (0-109) | 55 (0-85) | <0.001 |
| 0-20 | 70 (9%) | 66 (10%) | 4 (4%) | 0.067 |
| 21-40 | 278 (35%) | 263 (39%) | 15 (14%) | <0.001 |
| 41-60 | 315 (40%) | 272 (40%) | 43 (40%) | 1 |
| 60 | 124 (16%) | 79 (12%) | 45 (42%) | <0.001 |
| *Sex* |  |  |  | 0.205 |
| Male | 505 (64%) | 430 (63%) | 75 (70%) |  |
| Female | 282 (36%) | 250 (37%) | 32 (30%) |  |
| Health care workers | 53 (7%) | 52 (8%) | 1 (1%) | 0.006 |
| Patient presented at health facility | 524 (67%) | 418 (61%) | 106 (99%) | <0.001 |
| *Underlying comorbidity* | 340 (43%) | 267 (39%) | 73 (68%) | <0.001 |
| Cardiovascular disease | 130 (17%) | 98 (14%) | 32 (30%) | <0.001 |
| Diabetes | 116 (15%) | 87 (13%) | 29 (27%) | <0.001 |
| HIV | 53 (7%) | 42 (6%) | 11 (10%) | 0.172 |
| Cancer | 30 (4%) | 22 (3%) | 8 (7%) | 0.063 |
| Chronic Renal Disease | 24 (3%) | 14 (2%) | 10 (9%) | <0.001 |
| Chronic Obstructive Pulmonary Disease | 21 (3%) | 13 (2%) | 8 (7%) | 0.003 |
| *Symptoms at admission* |  |  |  |  |
| Present during admission | 455 (58%) | 376 (55%) | 79 (74%) | <0.001 |
| Cough | 337 (43%) | 284 (42%) | 53 (50%) | 0.160 |
| Loss of Taste or Smell | 279 (35%) | 208 (31%) | 71 (66%) | <0.001 |
| Fever | 126 (16%) | 108 (16%) | 18 (17%) | 0.917 |
| Headache | 99 (13%) | 94 (14%) | 5 (5%) | 0.013 |
| Muscle pains | 98 (12%) | 80 (9%) | 18 (17%) | 0.188 |
| Fatigue | 70 (9%) | 66 (10%) | 4 (4%) | 0.069 |
| Body weakness | 35 (4%) | 29 (4%) | 6 (6%) | 0.708 |
| Chest Pains | 26 (3%) | 23 (3%) | 3 (3%) | 0.984 |
| *Duration of onset of symptoms to:* |  |  |  |  |
| i) Hospital admission | 7 (0-53) | 7 (0-53) | 7 (0-38) | 0.846 |
| ii) ICU admission | 6 (0-38) | 7 (0-25) | 5 (0-38) | 0.282 |
| iii) Death | 16 (1-65) |  | 16 (1-65) | <0.001 |
| *Hospital course* |  |  |  |  |
| ICU admission | 90(11%) | 44 (6%) | 46 (43%) | <0.001 |
| Ventilation | 59(7%) | 13 (2%) | 46 (43%) | <0.001 |

More than half (58%) of the patients were reported to have clinical symptoms during admission, with cough (43%), loss of taste or smell (35%), fever (16%), headaches (13%) and muscle pains (12%) being the most frequent. 79 (11%) of patients required ICU admission and 59(7%) were mechanically ventilated. 107 (13.5%) of patients died. From the onset of symptoms, the average duration of hospital admission, ICU admission and death were 7, 6 and 16 days respectively. Table 1 provides a summary of the baseline characteristics of the patients included in the study.

### Laboratory findings of the patients

To determine the laboratory findings associated with survival of patients admitted with COVID-19, blood samples were collected, and tests carried out. The tests included haematology for 448 (57%) of the patients, creatinine (n=433, 55%), D-dimer (n=94, 12%), liver function test (n=421, 53%), C-reactive protein (n=184, 23%) and procalcitonin (n=14, 2%). A majority of patients analysed had neutropenia, lymphopenia, elevated aspartate aminotransferase, elevated lactate dehydrogenase and elevated C-reactive protein. The results showed differences in many of these parameters when compared between the survivors and the non-survivors (Table 2).

**Table 2:** **Laboratory results of the patients admitted in Kenyan health facilities with COVID-19**

| Parameter | All patients | Survivors | Non-survivors | P value |
| --- | --- | --- | --- | --- |
| Leucocyte count (*10<sup>9</sup> L; normal range 4-10) | *n*=448 | *n*=352 | *n*=96 |  |
| Increased | 99 (22%) | 55 (16%) | 44 (46%) | <0.001 |
| Segmented neutrophils (normal range 45-75%) |  | *n*=354 | *n*=94 |  |
| Decreased | 289 (65%) | 223 (63%) | 66 (70%) | <0.001 |
| Increased | 41 (11%) | 23 (6%) | 18 (19%) | 0.468 |
| Lymphocyte (normal range 25-40%) |  | *n*=355 | *n*=93 |  |
| Decreased | 309 (69%) | 227 (64%) | 82 (88%) | <0.001 |
| Increased | 60 (13%) | 58 (16%) | 2 (2%) | <0.001 |
| Haemoglobin (g/dL normal range: Male- 14-17, Female 12-16) | *n*=448 | *n*=349 | *n*=99 |  |
| Decreased | 128 (29%) | 76 (22%) | 52 (53%) | <0.001 |
| Aminotransferase, Alanine (U/L normal range: <35) | *n*=421 | *n*=316 | *n*=105 |  |
| Increased | 201 (48%) | 132 (42%) | 69 (66%) | <0.001 |
| Aminotransferase, Aspartate (U/L normal range: <35) | *n*=421 | *n*=316 | *n*=105 |  |
| Increased | 214 (51%) | 131 (41%) | 83 (79%) | <0.001 |
| Lactose dehydrogenase (U/L normal range: 60-100) | *n*=104 | *n*=66 | *n*=38 |  |
| Increased | 101 (97%) | 64 (97%) | 37 (97%) | 1 |
| Potassium (mmol/L normal range: 3.5-5) | *n*=433 | *n*=328 | *n*=105 |  |
| Decreased | 35 (8%) | 28 (9%) | 7 (7%) | 0.030 |
| Increased | 61 (15%) | 38 (12%) | 23 (22%) | 0.178 |
| C-reactive protein (mg/L normal range: <5) | *n*=197 | *n*=137 | *n*=60 |  |
| Increased | 155 (79%) | 96 (70%) | 59 (98%) | <0.001 |

### Radiological features

Severely ill patients got chest imaging (n=101). The most common findings included presence of ground glass opacities in 73(72%), local patchy shadowing in 59(58%), diffuse patchy shadowing in 61(60%) and interstitial abnormalities in 29(29%).

### Determinants of time-to death for COVID-19 patients admitted in health facilities

To determine the factors associated with death outcomes for hospitalized COVID-19 patients, we carried out Cox proportional hazard regression analysis. This was carried out in two stages: univariable analysis of all putative factors, followed by multivariable analysis to identify the significant factors associated with death outcomes for patients. Table 3 shows the results of the univariable analysis for all putative factors in the study dataset for COVID-19 mortality. Factors with a *p*-value of < 0.2 were offered to the multivariable analysis followed by model reduction.

**Table 3:** Univariable analysis of time to death for COVID-19 patients.

| Characteristic | Hazard Ratio | 95% CI | p-value |
| --- | --- | --- | --- |
| **Age group (*n*=787)** |  |  |  |
| 20 |  |  |  |
| 21-40 | 1.58 | 0.62, 4.06 | 0.3 |
| 41-60 | 3.66 | 1.49, 9.03 | 0.005 |
| 60 | 5.61 | 2.24, 14.0 | <0.001 |
| **Gender (*n*=787)** |  |  |  |
| Female |  |  |  |
| Male | 1.62 | 1.17, 2.24 | 0.004 |
| **Chronic conditions**<br>(n=787) |  |  |  |
| No |  |  |  |
| Yes | 2.62 | 1.92, 3.58 | <0.001 |
| **Ventilation** (n=787) |  |  |  |
| No |  |  |  |
| Yes | 12.3 | 9.06, 16.6 | <0.001 |
| **HIV** (n=787) |  |  |  |
| No |  |  |  |
| Yes | 1.49 | 0.83, 2.68 | 0.2 |
| **Reason hospitalization**<br>(n=787) |  |  |  |
| Other Reason |  |  |  |
| Visited Hospital | 106 | 14.7, 758 | <0.001 |
| **Chronic renal disease**<br>(n=787) |  |  |  |
| No |  |  |  |
| Yes | 2.77 | 1.80, 4.25 | <0.001 |
| **Asthma** (n=787) |  |  |  |
| No |  |  |  |
| Yes | 1.06 | 0.26, 4.26 | >0.9 |
| **Tuberculosis** (n=787) |  |  |  |
| No |  |  |  |
| Yes | 1.38 | 0.34, 5.58 | 0.6 |
| **Diabetes** (n=740) |  |  |  |
| No |  |  |  |
| Yes | 2.02 | 1.44, 2.85 | <0.001 |
| **Clinical symptoms**<br>(n=787) |  |  |  |
| No |  |  |  |
| Yes | 1.92 | 1.38, 2.67 | <0.001 |
| **Number of symptoms** | 1.13 | 1.04, 1.22 | 0.003 |
| **Headache** (*n*=787) |  |  |  |
| No |  |  |  |
| Yes | 0.33 | 0.18, 0.62 | <0.001 |
| **Fatigue** (*n*=787) |  |  |  |
| No |  |  |  |
| Yes | 0.53 | 0.31, 0.90 | 0.020 |
| **Myalgia or arthralgia**<br>(*n*=787) |  |  |  |
| No |  |  |  |
| Yes | 0.24 | 0.03, 1.71 | 0.2 |
| **Shortness of breath**<br>(*n*=787) |  |  |  |
| No |  |  |  |
| Yes | 1.97 | 1.34, 2.89 | <0.001 |
| **Fever** (*n*=787) |  |  |  |
| No |  |  |  |
| Yes | 1.92 | 1.39, 2.65 | <0.001 |
| **Cough** (*n*=787) |  |  |  |
| No |  |  |  |
| Yes | 1.25 | 0.93, 1.67 | 0.14 |
| **Sore throat** (*n*=787) |  |  |  |
| No |  |  |  |
| Yes | 1.86 | 0.92, 3.79 | 0.086 |
| **Weakness** (*n*=787) |  |  |  |
| No |  |  |  |
| Yes | 1.80 | 1.02, 3.16 | 0.042 |
| **Chest pains** (*n*=740) |  |  |  |
| No |  |  |  |
| Yes | 0.62 | 0.25, 1.50 | 0.3 |
| **Loss of taste or smell**<br>(*n*=787) |  |  |  |
| No |  |  |  |
| Yes | 3.43 | 2.52, 4.68 | <0.001 |
| **White blood cells**<br>(*n*=558) |  |  |  |
| Normal |  |  |  |
| Decreased | 0.57 | 0.26, 1.24 | 0.2 |
| Increased | 3.14 | 2.27, 4.34 | <0.001 |
| **Neutrophils** (*n*=559) |  |  |  |
| Normal |  |  |  |
| Decreased | 3.34 | 1.88, 5.95 | <0.001 |
| Increased | 6.13 | 3.22, 11.7 | <0.001 |
| **Lymphocytes** (*n*=558) |  |  |  |
| Normal |  |  |  |
| Decreased | 4.17 | 1.95, 8.90 | <0.001 |
| Increased | 0.30 | 0.06, 1.45 | 0.13 |
| **Haemoglobin** (*n*=554) |  |  |  |
| Normal |  |  |  |
| Decreased | 2.81 | 2.01, 3.92 | <0.001 |
| Increased | 0.88 | 0.40, 1.94 | 0.8 |
| **Alanine aminotrans-**<br>**ferase** (*n*=500) |  |  |  |
| Normal |  |  |  |
| Increased | 2.13 | 1.54, 2.95 | <0.001 |
| **Aspartate aminotrans-**<br>**ferase** (*n*=502) |  |  |  |
| Normal |  |  |  |
| Increased | 3.24 | 2.25, 4.68 | <0.001 |
| **Potassium** (*n*=539) |  |  |  |
| Normal |  |  |  |
| Decreased | 1.15 | 0.65, 2.05 | 0.6 |
| Increased | 1.72 | 1.20, 2.46 | 0.003 |
| **C reactive protein**<br>(*n*=263) |  |  |  |
| Normal |  |  |  |
| Increased | 25.9 | 3.61, 186 | 0.001 |

Figure 1 shows the association between the patient level factors and risk of COVID-19 mortality among the hospitalised patients. Risk of death increased with age with patients over 60 years of age having more than one and half increased risk of death compared to those below 60 years [HR 1.57 (1.13 – 2.19)]. Men had a higher risk of COVID-19 mortality compared to women, while those with at least one underlying co-morbidity had an increase in risk of death compared to those without. Presence of clinical symptoms was associated with increased risk of COVID-19 mortality.

**Figure 1:**
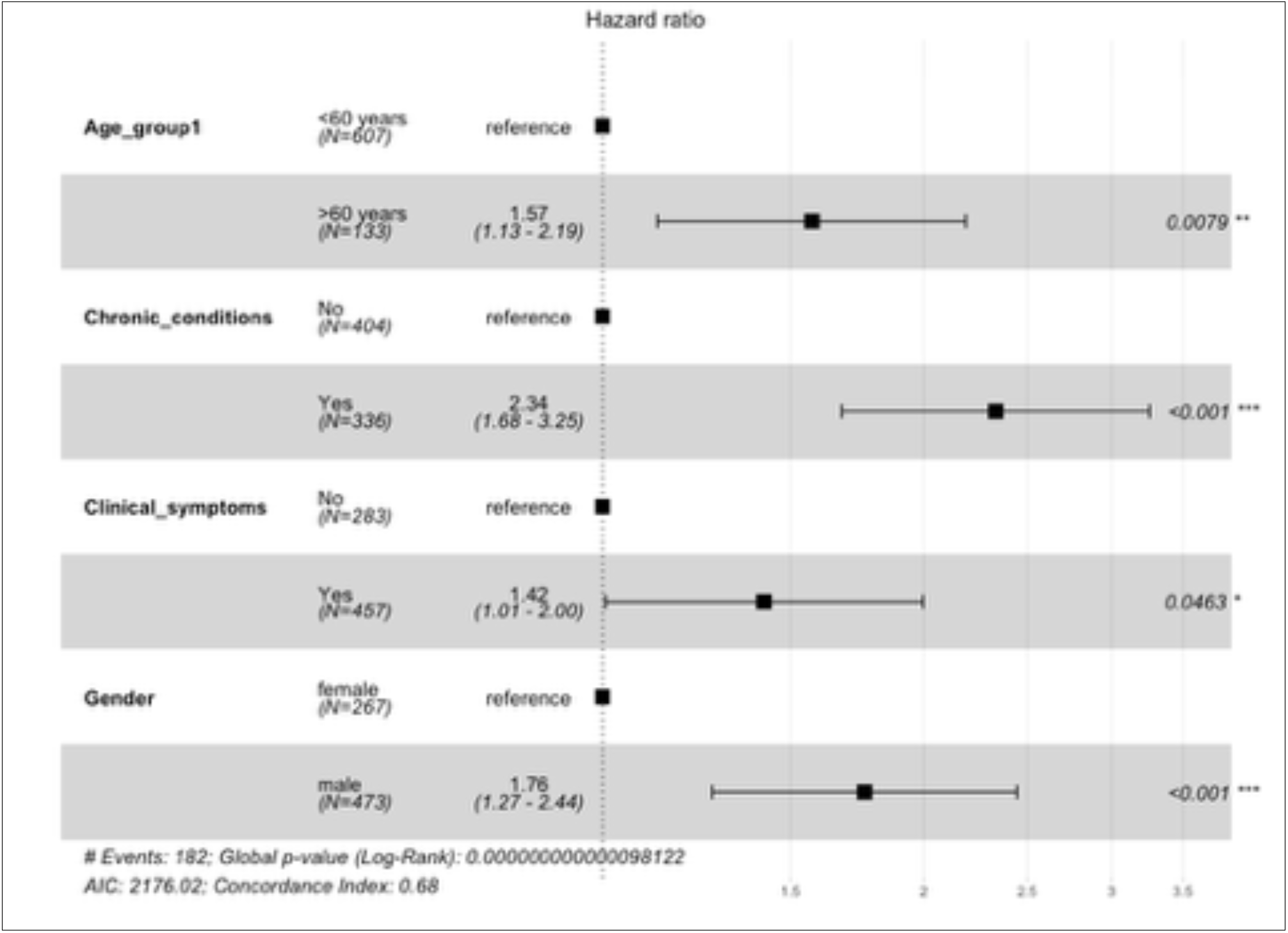
Figure showing the hazard ratios of the statistically significant factors in the multivariable model that are associated with death outcomes among COVID-19 patients admitted in the health facilities

Several laboratory test results were associated with higher risk of mortality (Figure 2). Increased white blood cell counts, both neutrophilic and neutropenia as well as lymphopenia, low haemoglobin and elevated liver enzymes were associated with increased risk of death from COVID-19.

**Figure 2:**
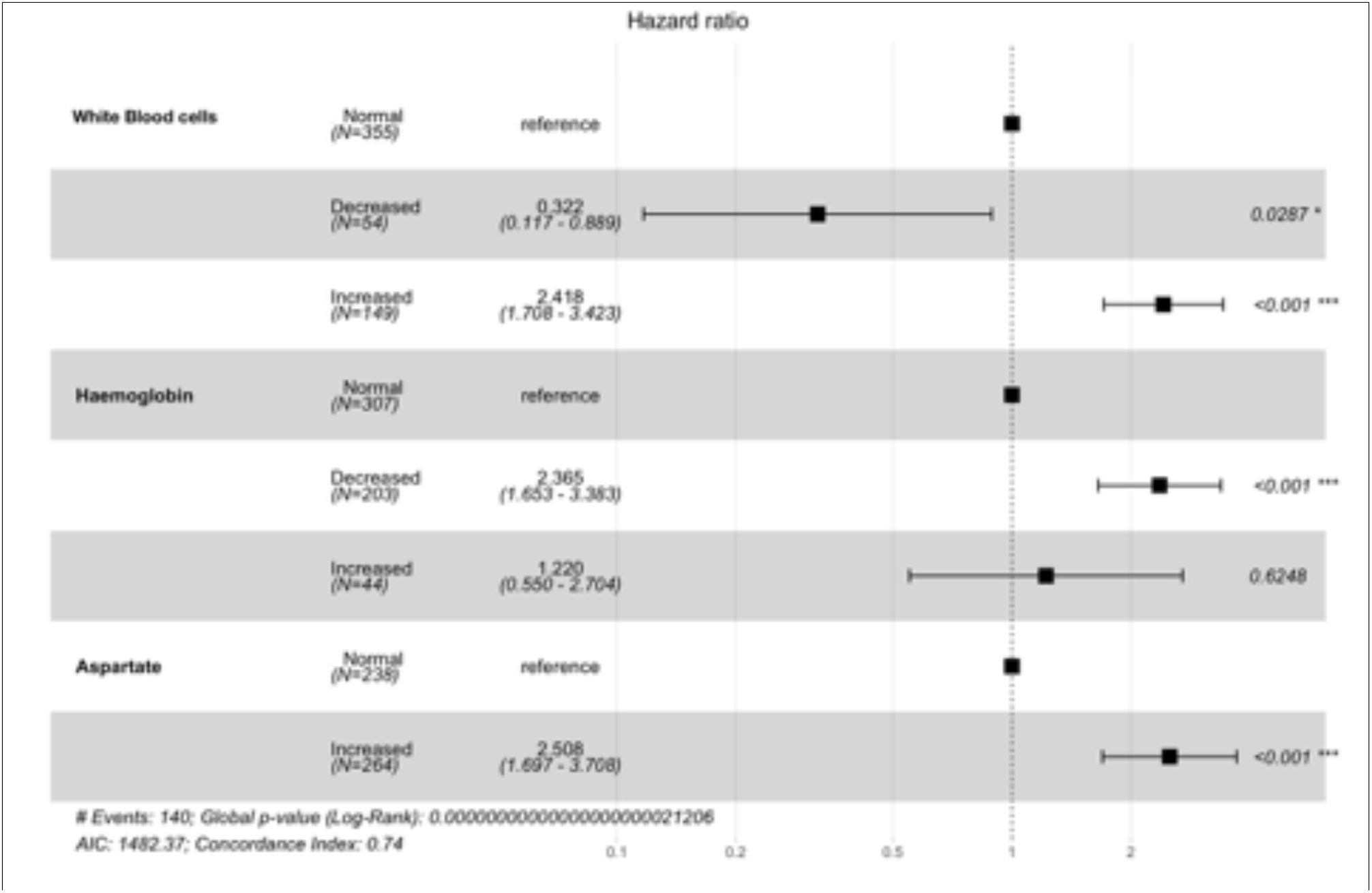
Figure showing the hazard ratios of the laboratory parameters from the multivariate model. An increase in aspartate, decrease in haemoglobin and an increase in the white blood cells increased the hazard ratio by more than 2.

## Discussion

This prospective, multi-centre study provides a summary of the epidemiological and clinical features of people with SARS-CoV-2 infection and COVID-19 of varying severity, and explores predictors of mortality.

Among COVID-19 positive patients admitted to six COVID-19 hospitals in Kenya between March - August 2020, the median age was 43 years, with 42% below the age of 40 and majority were male (64%). About 43% were found to have underlying chronic conditions, most commonly cardiovascular disease (17%), diabetes mellitus (15%), HIV (7%), malignancies (4%), chronic kidney disease (3%) and chronic airway disease (3%).

The incidence of ICU admission was 90(11%) with 59(7%) patients receiving mechanical ventilation. This incidence of severe illness requiring ICU admission is lower than that observed in other cohorts largely from developing countries. Cummings et al report on a New York cohort in which the incidence of ICU admission was 22%. [13]

There are several factors that may account for this lower disease severity. The initial Kenya national containment strategy included admission of all persons testing COVID-19 positive even in the absence of symptoms, this was at a time when over 80% of the patients were asymptomatic [14]. Our patient population was also younger than that reported elsewhere, with a median age of the general population of 20 years compared to 45 in Italy and 44.9 in Spain, 38.2 in USA and 33 in Brazil, all countries that have seen significantly higher morbidity and mortality. In the New York cohort, they reported a mean age of 62 years and a high prevalence of hypertension (63%) in patients admitted to the critical care units [13]. The prevalence of underlying chronic conditions was lower in our cohort than that reported elsewhere. De Souza and others in an analysis of the epidemic in Brazil, report prevalence of 66.5% of cardiovascular disease and 54.5% diabetes in patients with COVID-19 [15]. Older age and presence of underlying co-morbidities have both been associated with increased risk of severe outcomes in COVID-19 [16,17].

58% of our patients were symptomatic at the time of admission with the most common symptoms being cough (58%), loss of taste and smell (35%), and fever (16%). Earlier reports on the epidemic reported fever as the most common symptom followed by cough. In a systematic review and meta-analysis, Hu found the prevalence of fever to be 85.6%, cough at 65.7%, other common symptoms were fatigue and dyspnoea [18]. Loss of smell and taste was fairly prevalent in our population as has been reported elsewhere [19].

Non-survivors were more likely to be older, have an underlying comorbidity with cardiovascular disease, diabetes mellitus, renal insufficiency and chronic obstructive airway disease more likely to be present in non-survivors. This is in keeping with reports from other cohorts [20]. Many chronic diseases may lead to a state of heightened inflammation and impaired immune responses with an overall lowering of immunity.

On cox proportional hazard regression analysis, we found significantly increased risk of death with older age (>60), male gender and in patients with co-morbidities. Men with COVID-19 have been shown to be at higher risk for worse outcomes and mortality irrespective of age [21]. Possible explanations for this include the higher prevalence of high-risk behaviour including smoking and attendant lung injury, higher prevalence of underlying co-morbidities and other yet to be fully defined biologic differences.

We found that the presence of a comorbidity was associated with increased mortality, HR 2.34 (CI 1.69-3.25). Co-morbidities have been associated with higher risk of severe outcomes in many populations. Data from China showed that the hazard ratio of severe outcomes including admission to ICU, invasive ventilation and death was 1.79 for patients with at least one comorbidity and 2.59 for patients with two or more co-morbidities [17].

As countries think of strategies to reduce disease transmission and reduce risk of severe disease and mortality, it is important that these risk factors of older age and presence of co-morbidity are taken in to account and strategies that identify and shield those at highest risk as defined here are adopted.

We found various laboratory parameters to be associated with increased risk of death, these included leucocytosis, lymphopaenia, transaminitis and elevated CRP. Lymphopaenia has been shown to occur frequently in patients with COVID-19 and to predict severe disease [22]. Lymphopaenia may result either from suppression of the bone marrow, direct infection and destruction or a cytotoxic mediated killing of lymphocytes. A functional exhaustion of antiviral lymphocytes has also been reported [23].

We found elevated alanine and aspartate aminotransferase in 48% and 51% of patients respectively. Elevations in liver enzymes is common and has been reported to range from 16-53% in various studies [24,25]. Boregowda et al in a meta-analysis of studies comparing liver chemistries in mild and severe disease, showed that elevated liver enzymes were associated with severe disease, and predicted mortality [25]. This finding is further strengthened by our our study where the presence of elevated aspartate aminotransferase was associated with a hazard ratio of death of 2.5 (CI 1.69-3.7)

This main strengths of this study is that it was multi-center and included asymptomatic and mild cases, which provides a more comprehensive analysis of the presentation of COVID-19 and reduces bias. Limitations of this study include the absence of laboratory parameters for some of the study patients, pulse oximetry was not routinely recorded during the initial period of the outbreak and we did not have access to other laboratory markers that have been shown to predict mortality including D-dimers and interleukin 6.

## Conclusion

In conclusion, this study demonstrates that patients with COVID-19 in Kenya were fairly young with a low rate of severe disease. Age >60, male gender, presence of co-morbidities, leucocytosis, lymphopaenia and elevated transaminases predicted mortality.

## Data Availability

De-identified participant data is available and will be made publicly available 6 months after publication of this manuscript or earlier for related work

https://osf.io/b6wkr/

## References

1. WHO Coronavirus Disease (COVID-19) Dashboard | WHO Coronavirus Disease (COVID-19) Dashboard [Internet]. [cited 2020 Oct 26]. Available from: https://covid19.who.int/

2. Guan W, Ni Z, Hu Y et al. Clinical characteristics of Corona Virus Disease 2019 in China. N Engl J Med 2020; 382: 1708–1720

3. Nachega JB, Ishoro DK, Otokoye JO et al. Clinical Characteristics and Outcomes of Pa- tients Hospitalized for COVID-19 in Africa: Early Insights from the Democratic Republic of the Congo. Am J Trop. Med. Hyg., 00(0), 2020, pp 1–10

4. Bruce Aylward (WHO); Wannian Liang (PRC). Report of the WHO-China Joint Mission on Coronavirus Disease 2019 (COVID-19). Vol. 1, The WHO-China Joint Mission on Coronavirus Disease 2019. 2020. p. 40.

5. Wu Z, McGoogan JM. Characteristics of and Important Lessons from the Coronavirus Disease 2019 (COVID-19) Outbreak in China: Summary of a Report of 72314 Cases from the Chinese Center for Disease Control and Prevention. JAMA - J Am Med Assoc. 2020 Apr 7;323(13):1239–42.

6. Wang D, Hu B, Hu C, et al. Clinical Characteristics of 138 Hospitalized Patients with 2019 Novel Coronavirus-Infected Pneumonia in Wuhan, China. JAMA - J Am Med Assoc. 2020 Mar 17;323(11):1061–9.

7. Rodriguez-Morales AJ, Cardona-Ospina JA, Gutiérrez-Ocampo E, et al. Clinical, laboratory and imaging features of COVID-19: A systematic review and meta-analysis. Travel Medicine and Infectious Disease. 2020.

8. Onder G, Rezza G, Brusaferro S. Case-Fatality Rate and Characteristics of Patients Dying in Relation to COVID-19 in Italy. JAMA - Journal of the American Medical Association. American Medical Association; 2020.

9. Korea Centers for Disease Control. Updates on COVID-19 in Korea. 2020;

10. Stokes EK, Zambrano LD, Anderson KN, et al. Coronavirus Disease 2019 Case Sur- veillance - United States, January 22-May 30, 2020. MMWR Morb Mortal Wkly Rep. 2020;

11. Wu C, Chen X, Cai Y, Xia J, et al. Risk Factors Associated with Acute Respiratory Distress Syndrome and Death in Patients with Coronavirus Disease 2019 Pneumonia in Wuhan, China. JAMA Intern Med. 2020;

12. R Core Team. R: A language and environment for statistical computing. Vienna, Austria; 2013.

13. Cummings MJ, Baldwin MR, Abrams D, et al. Epidemiology, clinical course, and outcomes of critically ill adults with COVID-19 in New York City: a prospective cohort study. Lancet. 2020 Jun 6;395(10239):1763–70.

14. Ministry of Health Emergency Operations Centre. COVID-19 Outbreak in Kenya Daily Situation Reports. Nairobi; 2020.

15. De Souza WM, Buss LF, Candido D da S, et al. Epidemiological and clinical characteristics of the COVID-19 epidemic in Brazil. Nat Hum Behav. 2020;4(8):856–65.

16. Shahid Z, Kalayanamitra R, McClafferty B, et al. COVID-19 and older adults: what we know. J Am Geriatr Soc. 2020;68(5):926–9.

17. Guan W, Liang W, Zhao Y, et al. Comorbidity and its impact on 1590 patients with COVID-19 in China: a nationwide analysis. Eur Respir J. 2020 May 1;55(5):2000547.

18. Hu Y, Sun J, Dai Z, et al. Prevalence and severity of corona virus disease 2019 (COVID-19): A systematic review and meta-analysis. Vol. 127, Journal of Clinical Virology. Elsevier B.V.; 2020.

19. Meng X, Deng Y, Dai Z, Meng Z. COVID-19 and anosmia: A review based on up-to-date knowledge. Vol. 41, American Journal of Otolaryngology - Head and Neck Medicine and Surgery. W.B. Saunders; 2020. p. 102581.

20. Yang J, Zheng Y, Gou X, et al. Prevalence of comorbidities and its effects in coronavirus disease 2019 patients: A systematic review and meta-analysis. Int J Infect Dis. 2020 May 1;94:91–5.

21. Jin JM, Bai P, He W, et al. Gender Differences in Patients With COVID-19: Focus on Severity and Mortality. Front Public Heal. 2020 Apr 29;8:152.

22. Zhao Q, Meng M, Kumar R, et al. Lymphopenia is associated with severe coronavirus disease 2019 (COVID-19) infections: A systemic review and meta-analysis. Int J Infect Dis. 2020/05/04. 2020 Jul;96:131–5.

23. Zheng M, Gao Y, Wang G, et al. Functional exhaustion of antiviral lymphocytes in COVID-19 patients. Vol. 17, Cellular and Molecular Immunology. Springer Nature; 2020. p. 533–5.

24. Xie H, Zhao J, Lian N, Lin S, Xie Q, Zhuo H. Clinical characteristics of non-ICU hospitalized patients with coronavirus disease 2019 and liver injury: A retrospective study. Liver Int. 2020 Jun 1;40(6):1321–6.

25. Boregowda U, Aloysius MM, Perisetti A, Gajendran M, Bansal P, Goyal H. Serum Activity of Liver Enzymes Is Associated With Higher Mortality in COVID-19: A Systematic Review and Meta-Analysis. Front Med. 2020 Jul 22;7:431.

